# Mixtures of multiple air pollutants from specific industrial or residential sources and mortality from ischemic heart disease, cardiovascular disease, and non-accidental causes: A large general population Canadian cohort study

**DOI:** 10.1101/2025.01.07.25320163

**Authors:** Naizhuo Zhao, Toyib Olaniyan, Kimberly Mitchell, Mathieu Rouleau, Ivana Popadic, Angelos Anastasopolos, Michael Tjepkema, Sabit Cakmak

## Abstract

**Background:** Previous studies on ambient air pollution and mortality typically focus on individual pollutants rather than their mixtures, and overall pollution rather than air pollution from specific sources. We aimed to assess the associations between ambient mixtures of fine particulate matter (PM_2.5_), sulfur dioxide (SO_2_), nitrogen dioxide (NO_2_), and ozone (O_3_) from various industrial and residential sources, and deaths from ischemic heart disease (IHD), cardiovascular disease (CVD) and non-accidental causes.

**Methods:** We linked the 2006 Canadian Census Health and Environment Cohort (CanCHEC) with the Canadian Vital Statistics Database, identifying 56190, 98185, and 381050 deaths between 2006 and 2019 from IHD, CVD, and non-accidental causes, respectively. Annual average concentrations of PM_2.5_, SO_2_, NO_2_, and O_3_ from upstream petroleum, downstream petroleum, non-ferrous smelting, chemical industry and residential fuel combustion were estimated using the Global Environmental Multiscale-Modelling Air Quality and Chemistry (GEM-MACH) model. These concentrations were assigned to CanCHEC participants based on their annual residential postal codes. Quantile g-computation models were used to calculate hazard ratios (HRs) for deaths from IHD, CVD, and non-accidental causes per quartile increase in all four air pollutants from each specific sector.

**Results:** We observed significant associations between the mixture of air pollutants and deaths from IHD, CVD, and non-accidental causes for emissions from upstream petroleum [HR: 1.18 (95% CI: 1.12-1.24), 1.12 (1.08-1.16), and 1.05 (1.04-1.05)], downstream petroleum [1.06 (1.05-, 1.04 (1.03-1.05), and 1.03 (1.02-1.03)], the chemical industry [1.10 (1.08-1.13), 1.07 (1.06-1.09), and 1.10 (1.08-1.13)], and residential fuel combustion [1.18 (1.12-1.23), 1.12 (1.08-1.16), and 1.07 (1.05-1.09)]. PM_2.5_ and SO_2_ contributed more to the increased risk of death than NO_2_ and O_3_. Mortality from CVD or non-accidental causes was not associated with the mixtures of air pollutants from non-ferrous smelting.

**Conclusions:** Ambient PM_2.5_ and SO_2_ from certain sectors, but not all, greatly contribute to the increased risk of non-accidental, IHD, and CVD deaths.

## Introduction

A large body of epidemiology studies have shown that long-term exposure to air pollution increases the risk for non-accidental mortality and specific cardiovascular diseases (CVD) such as ischemic heart disease (IHD).^1–3^ However, most of the studies have focused on the associations between premature deaths and exposure to individual air pollutants, overlooking the fact that we are typically exposed to multiple air pollutants such as fine particulate matter (PM_2.5_), sulfur dioxide (SO_2_), nitrogen dioxide (NO_2_), and ozone (O_3_) simultaneously. Recently, some studies have begun considering multiple air pollutants but they often rely on traditional statistical regression methods like multi-pollutant Cox proportional hazard models.^4,5^ These models fail to address the issue of collinearity caused by the inter-correlated air pollutant concentrations, potentially leading to inaccurate estimates or difficult to interpret results.^6,7^ Moreover, for aggregate pollutants such as PM_2.5_, the chemical composition of a given ambient concentration can vary considerably, reflecting emission profile differences between sources or sectors, which may lead to differences in its toxicity and health effects.^8,9^

Identifying the air pollutants from specific sources that pose the greatest harm to human health is a worthwhile endeavor. This knowledge allows air quality management strategies to prioritize the most toxic air pollutants and effectively enhance the health benefits of improved air quality. However, investigating the health effects of exposure to multiple air pollutants, particularly those emitted from the same source type, presents methodological challenges. Specialized statistical methods are required to avoid occurrence of collinearity.^6,7,10^

Quantile g-computation is a statistical model developed specifically to study combined health effects of multiple exposures.^10^ Quantile g-computation avoids collinearity by transforming continuous exposure variables to quantile scales, and uses index weights to quantify the relative contribution of each exposure on overall health effects. Furthermore, quantile g-computation has excellent computational efficiency, making it particularly suitable for analyzing large population cohorts with many covariates.^11^

Currently, few air quality modelling-based epidemiological studies have investigated the links between health effects and source-specific air pollutants other than for the transportation sector, notably in the Canadian context.^12–14^ A limited number of recent United States-based studies have estimated source-specific PM_2.5_ CV-related health risk for non-traffic source types by combining source apportionment modelling methods with epidemiological analysis.^15–18^

However, prior source apportionment-based epidemiological studies are typically limited to single pollutant analysis rather than examining mixtures of sector-specific particle and gaseous pollutants. Accordingly, the objectives of this study are to assess the associations between mortality from IHD, CVD, and non-accidental causes with mixtures of PM_2.5_, SO_2_, NO_2_, and O_3_ from five industrial and residential sectors including upstream petroleum, downstream petroleum, non-ferrous smelting, the chemical industry and residential fuel combustion. Additionally, this study aims to identify the individual air pollutants within these mixtures that may be associated with comparably greater health risk by using quantile g-computation models. To do this, we used a large Canadian general population cohort and a complex chemical transport model to estimate levels of the four air pollutants from the five specific sectors.

## Methods

### Study cohort and mortality outcomes

Our analyses were based on the 2006 Canadian Census Health and Environment Cohort (CanCHEC), which includes nearly 4 million people, representing approximately 20% of Canadian households enumerated in the 2006 long-form census.^19^ The cohort consists of non-institutionalized respondents of the long-form census questionnaire who consented to record linkage with their tax files. Postal codes of these CanCHEC participants, covering 2006 to 2019 were obtained from their longitudinal tax files. Details regarding the tax file linkage and imputation of missing postal codes have been described by Pappin et al^20^ and Zhao et al^21^. From the 2006 CanCHEC, we further selected individuals who were at least 25 years old on the census date (May 16, 2006), had lived in Canada for at least ten years prior to cohort entry, and held valid residential (non-business) postal codes. These criteria resulted in 3,019,125 individuals being included in our analyses.

The CanCHEC participants were also linked to the Canadian Vital Statistics Database and followed up from the census date to death or end of study (i.e. December 31, 2019). In the database, each death event was coded by its primary cause. The World Health Organization International Classification of Diseases-10th Revision codes were used to determine events (and their corresponding years) for IHD-specific (I20-I25), CVD-specific (I20–I25, I30–I51, I60–I69, I70), and all non-accidental mortalities (A-R).

### Exposure variables

Source/sector-specific annual average concentrations of ambient PM_2.5_, SO_2_, NO_2_, and O_3_ for 2015 and 2019 were modelled at a horizontal grid resolution of 10 km by 10 km using Environment and Climate Change Canada’s Global Environmental Multi-scale - Modelling air quality and Chemistry (GEM-MACH) model. GEM-MACH is a chemical transport model that integrates atmospheric chemistry and meteorological processes in-step. The GEM-MACH model incorporates inventoried air pollutant emissions from all point and area sources comprising a given sector and uses meteorological fields from a regional model to calculate the ambient concentrations of pollutants of interest. Outputs of the GEM-MACH model are compared with air quality surface observations from Canadian regional networks and the U.S. EPA’s AIRNow observation network to generate the estimates-of-error statistics. These errors are mostly correlated with terrain (e.g. land, water, and mountain) and thus, the estimates of the GEM-MACH model are further adjusted by terrain to more closely approximate real ground-level pollutant concentrations. More details regarding the GEM-MACH model and its application for estimating concentrations of air pollutants have been described by Gong et al^22^ and Whaley et al^23,24^.

To assess the relative contribution of a given sector to total population air pollution exposure, we first ran a GEM-MACH base case for 2015 and 2019 that included all emission sources. Then, we ran model simulations excluding emissions from that specific sector for the same year. The relative contributions of those specific sources were calculated by comparing the base case with the scenario in which all emissions from that interested sector were set to zero.

Our analysis revealed that the percentage contributions of residential fuel combustion, upstream and downstream petroleum, non-ferrous smelting, and the chemical industry to overall ambient air pollution remained relatively stable between 2015 and 2019. Using 2015 as the reference year, we applied these sector-specific percentage contributions to the total annual averages of ambient air pollution measurements from National Ambient Air Pollution Surveillance (NAPS) monitors to estimate source-specific air pollution concentrations from 2006 to 2019. As described by Pappin et al^20^, three-year moving averages with one-year lag exposures were assigned to the 3,019,125 individuals based on their annual residential postal codes. Descriptions of the specific air pollutant emission source types modelled within each sector are provided in Table S1.

### Covariates

The 2006 CanCHEC includes common socio-demographic factors such as age, sex, education level, household income, occupational class, marital status, labor force status, visible minority, indigenous identity, and immigrant status. These individual-level variables, collected at cohort entry, as part of the long-form census, were used as covariates in this study.

The Canadian Marginalization Index (CAN-Marg) is a set of area-based measures that quantifies socioeconomic inequality using four indicators: residential instability, material deprivation, dependency, and ethnic concentration.^25^ CAN-Marg was created from census data at the dissemination-area (DA) level, the smallest available standard geographic area.

Socioeconomic inequality in Canada, as captured by CAN-Marg, has been shown to be associated with health outcomes^25^, and thus its four sub-indexes were used as contextual covariates in this study.

To capture urban-rural distinctions and control for urban-rural differences in mortality rates, we included two more covariates: census metropolitan areas or census agglomerations (CMA/CA) size and urban form. CMA/CA size was categorized into five levels based on population (see Table 1 for the specific population size in each level).^26^ Urban form was created by combining population density and the most frequently used transportation mode, resulting in five categories: active urban core, transit-reliant suburb, car-reliant suburb, exurban, and non-CMA/CA.^27^

**Table 1.**
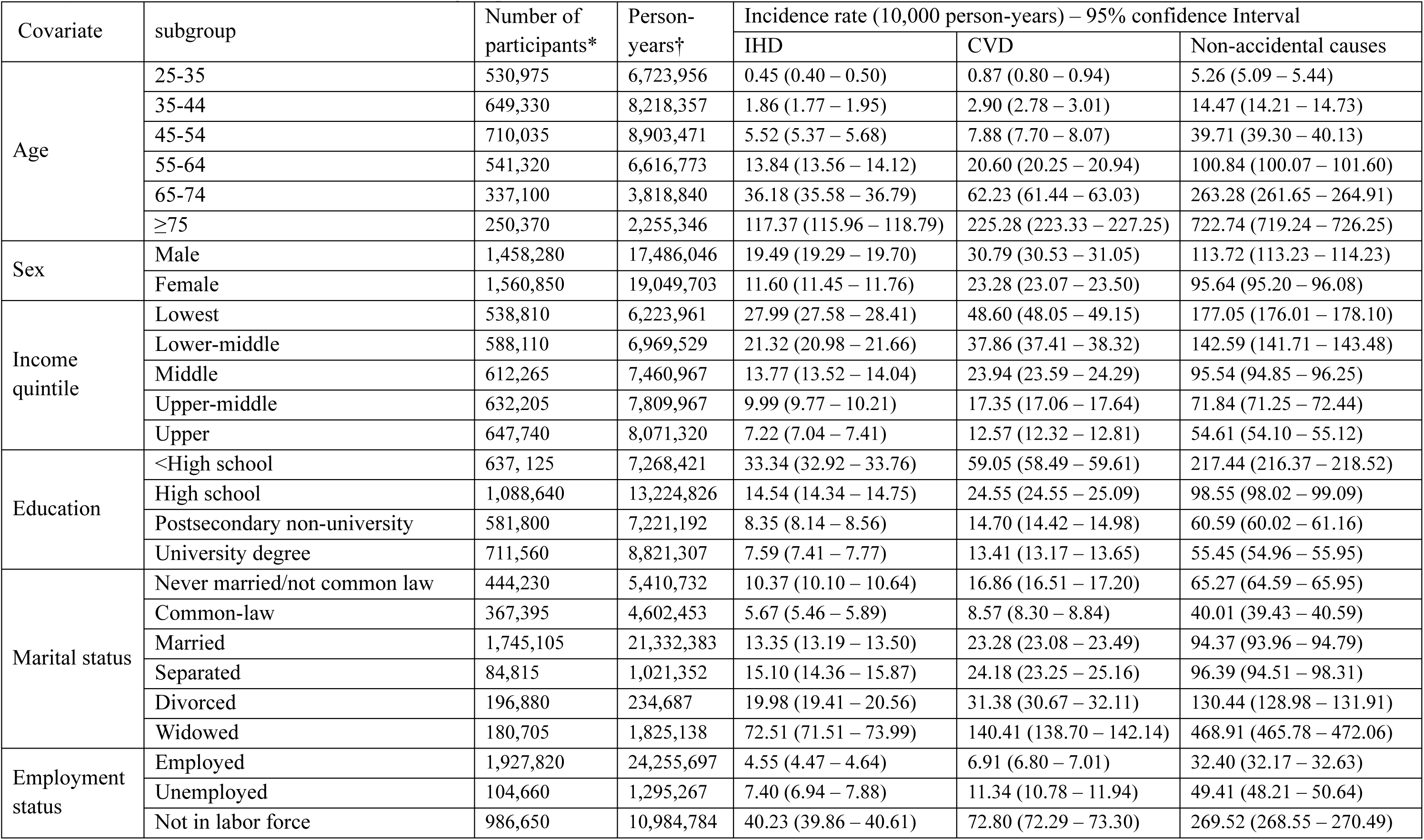
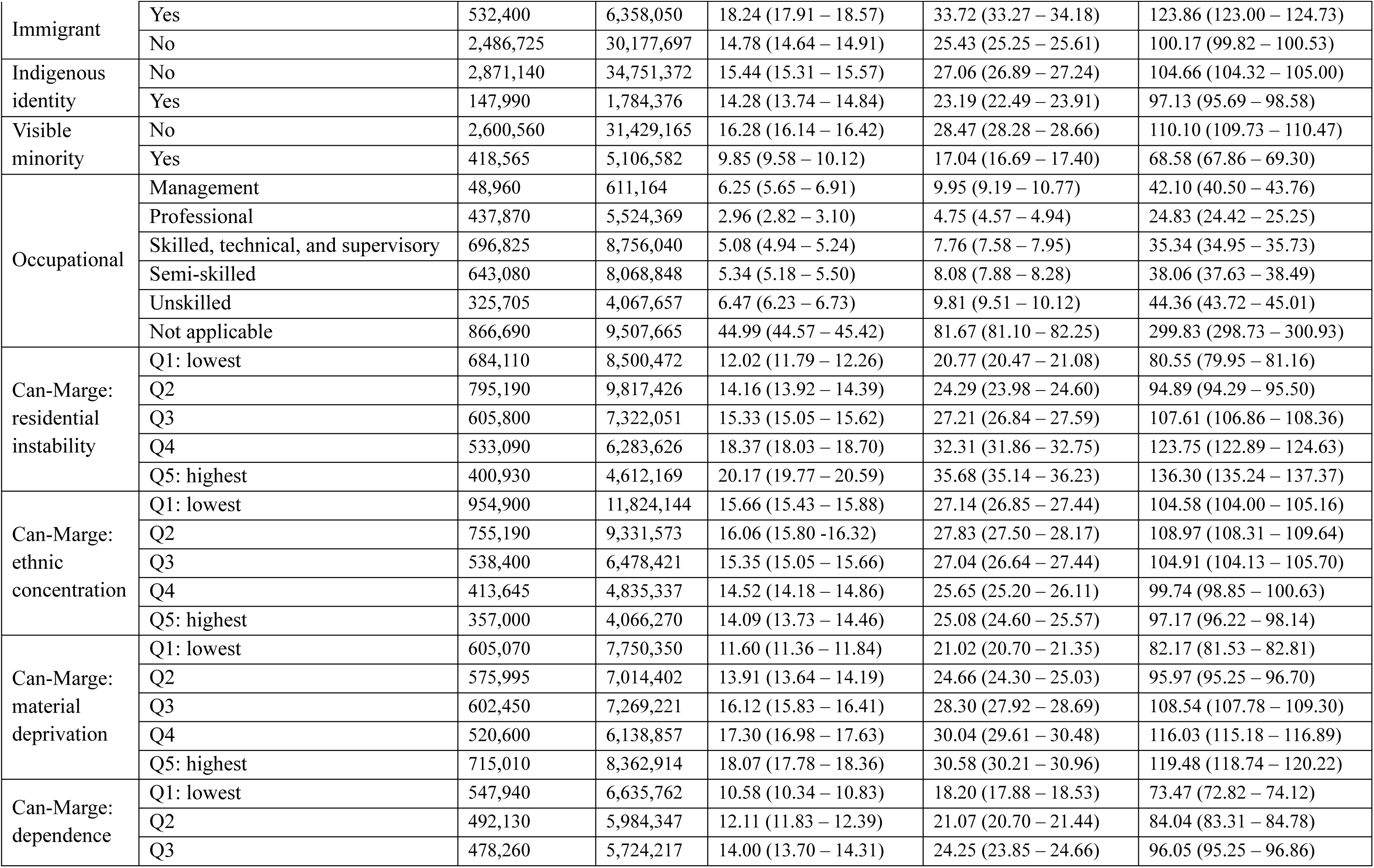
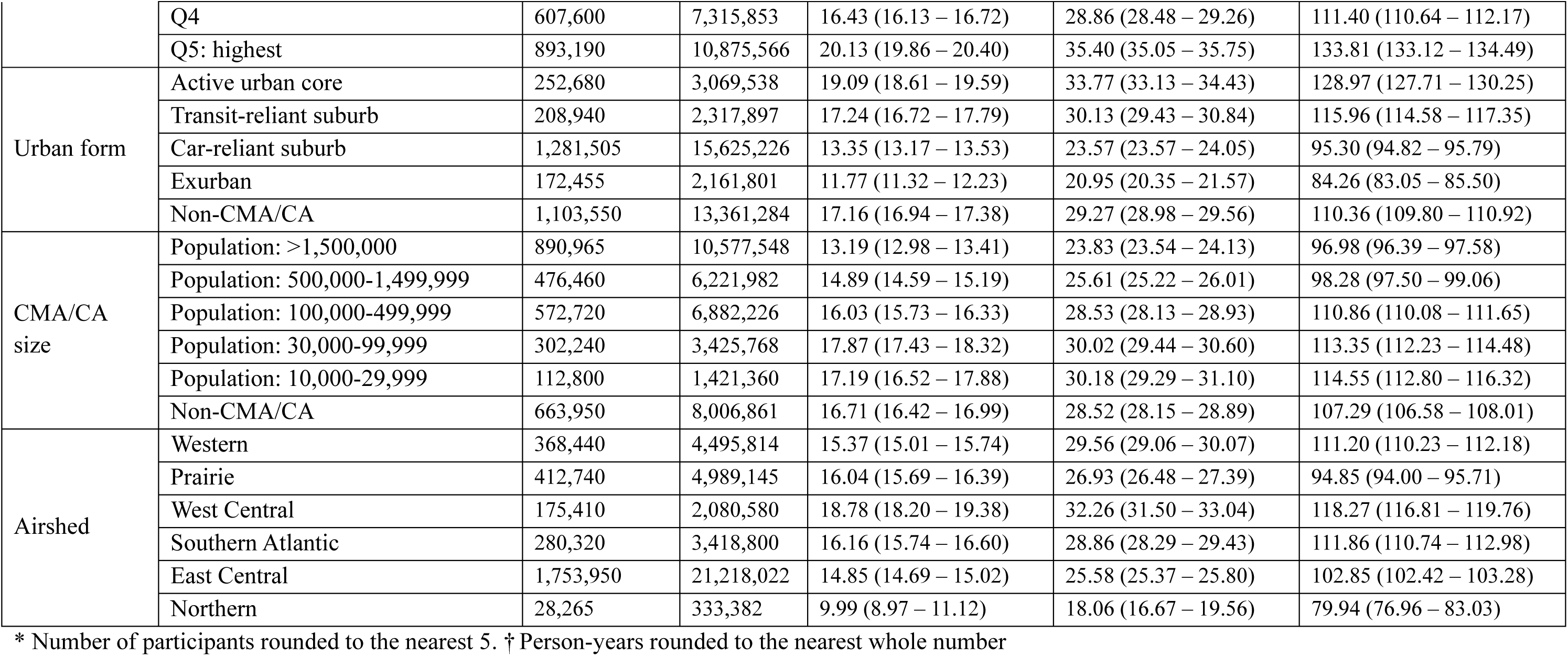
The number of subjects and incidence rates of deaths attributable from ischemic heart disease (IHD), cardiovascular disease (CVD), and non-accidental causes in each subgroup of each covariate.

| Covariate | subgroup | Number of participants* | Person-years† | Incidence rate (10,000 person-years) – 95% confidence Interval |  |  |
| --- | --- | --- | --- | --- | --- | --- |
|  |  |  |  | IHD | CVD | Non-accidental causes |
| Age | 25-35 | 530,975 | 6,723,956 | 0.45 (0.40 – 0.50) | 0.87 (0.80 – 0.94) | 5.26 (5.09 – 5.44) |
|  | 35-44 | 649,330 | 8,218,357 | 1.86 (1.77 – 1.95) | 2.90 (2.78 – 3.01) | 14.47 (14.21 – 14.73) |
|  | 45-54 | 710,035 | 8,903,471 | 5.52 (5.37 – 5.68) | 7.88 (7.70 – 8.07) | 39.71 (39.30 – 40.13) |
|  | 55-64 | 541,320 | 6,616,773 | 13.84 (13.56 – 14.12) | 20.60 (20.25 – 20.94) | 100.84 (100.07 – 101.60) |
|  | 65-74 | 337,100 | 3,818,840 | 36.18 (35.58 – 36.79) | 62.23 (61.44 – 63.03) | 263.28 (261.65 – 264.91) |
|  | ≥75 | 250,370 | 2,255,346 | 117.37 (115.96 – 118.79) | 225.28 (223.33 – 227.25) | 722.74 (719.24 – 726.25) |
| Sex | Male | 1,458,280 | 17,486,046 | 19.49 (19.29 – 19.70) | 30.79 (30.53 – 31.05) | 113.72 (113.23 – 114.23) |
|  | Female | 1,560,850 | 19,049,703 | 11.60 (11.45 – 11.76) | 23.28 (23.07 – 23.50) | 95.64 (95.20 – 96.08) |
| Income quintile | Lowest | 538,810 | 6,223,961 | 27.99 (27.58 – 28.41) | 48.60 (48.05 – 49.15) | 177.05 (176.01 – 178.10) |
|  | Lower-middle | 588,110 | 6,969,529 | 21.32 (20.98 – 21.66) | 37.86 (37.41 – 38.32) | 142.59 (141.71 – 143.48) |
|  | Middle | 612,265 | 7,460,967 | 13.77 (13.52 – 14.04) | 23.94 (23.59 – 24.29) | 95.54 (94.85 – 96.25) |
|  | Upper-middle | 632,205 | 7,809,967 | 9.99 (9.77 – 10.21) | 17.35 (17.06 – 17.64) | 71.84 (71.25 – 72.44) |
|  | Upper | 647,740 | 8,071,320 | 7.22 (7.04 – 7.41) | 12.57 (12.32 – 12.81) | 54.61 (54.10 – 55.12) |
| Education | <High school | 637,125 | 7,268,421 | 33.34 (32.92 – 33.76) | 59.05 (58.49 – 59.61) | 217.44 (216.37 – 218.52) |
|  | High school | 1,088,640 | 13,224,826 | 14.54 (14.34 – 14.75) | 24.55 (24.55 – 25.09) | 98.55 (98.02 – 99.09) |
|  | Postsecondary non-university | 581,800 | 7,221,192 | 8.35 (8.14 – 8.56) | 14.70 (14.42 – 14.98) | 60.59 (60.02 – 61.16) |
|  | University degree | 711,560 | 8,821,307 | 7.59 (7.41 – 7.77) | 13.41 (13.17 – 13.65) | 55.45 (54.96 – 55.95) |
| Marital status | Never married/not common law | 444,230 | 5,410,732 | 10.37 (10.10 – 10.64) | 16.86 (16.51 – 17.20) | 65.27 (64.59 – 65.95) |
|  | Common-law | 367,395 | 4,602,453 | 5.67 (5.46 – 5.89) | 8.57 (8.30 – 8.84) | 40.01 (39.43 – 40.59) |
|  | Married | 1,745,105 | 21,332,383 | 13.35 (13.19 – 13.50) | 23.28 (23.08 – 23.49) | 94.37 (93.96 – 94.79) |
|  | Separated | 84,815 | 1,021,352 | 15.10 (14.36 – 15.87) | 24.18 (23.25 – 25.16) | 96.39 (94.51 – 98.31) |
|  | Divorced | 196,880 | 234,687 | 19.98 (19.41 – 20.56) | 31.38 (30.67 – 32.11) | 130.44 (128.98 – 131.91) |
|  | Widowed | 180,705 | 1,825,138 | 72.51 (71.51 – 73.99) | 140.41 (138.70 – 142.14) | 468.91 (465.78 – 472.06) |
| Employment status | Employed | 1,927,820 | 24,255,697 | 4.55 (4.47 – 4.64) | 6.91 (6.80 – 7.01) | 32.40 (32.17 – 32.63) |
|  | Unemployed | 104,660 | 1,295,267 | 7.40 (6.94 – 7.88) | 11.34 (10.78 – 11.94) | 49.41 (48.21 – 50.64) |
|  | Not in labor force | 986,650 | 10,984,784 | 40.23 (39.86 – 40.61) | 72.80 (72.29 – 73.30) | 269.52 (268.55 – 270.49) |
| Immigrant | Yes | 532,400 | 6,358,050 | 18.24 (17.91 – 18.57) | 33.72 (33.27 – 34.18) | 123.86 (123.00 – 124.73) |
|  | No | 2,486,725 | 30,177,697 | 14.78 (14.64 – 14.91) | 25.43 (25.25 – 25.61) | 100.17 (99.82 – 100.53) |
| Indigenous identity | No | 2,871,140 | 34,751,372 | 15.44 (15.31 – 15.57) | 27.06 (26.89 – 27.24) | 104.66 (104.32 – 105.00) |
|  | Yes | 147,990 | 1,784,376 | 14.28 (13.74 – 14.84) | 23.19 (22.49 – 23.91) | 97.13 (95.69 – 98.58) |
| Visible minority | No | 2,600,560 | 31,429,165 | 16.28 (16.14 – 16.42) | 28.47 (28.28 – 28.66) | 110.10 (109.73 – 110.47) |
|  | Yes | 418,565 | 5,106,582 | 9.85 (9.58 – 10.12) | 17.04 (16.69 – 17.40) | 68.58 (67.86 – 69.30) |
| Occupational | Management | 48,960 | 611,164 | 6.25 (5.65 – 6.91) | 9.95 (9.19 – 10.77) | 42.10 (40.50 – 43.76) |
|  | Professional | 437,870 | 5,524,369 | 2.96 (2.82 – 3.10) | 4.75 (4.57 – 4.94) | 24.83 (24.42 – 25.25) |
|  | Skilled, technical, and supervisory | 696,825 | 8,756,040 | 5.08 (4.94 – 5.24) | 7.76 (7.58 – 7.95) | 35.34 (34.95 – 35.73) |
|  | Semi-skilled | 643,080 | 8,068,848 | 5.34 (5.18 – 5.50) | 8.08 (7.88 – 8.28) | 38.06 (37.63 – 38.49) |
|  | Unskilled | 325,705 | 4,067,657 | 6.47 (6.23 – 6.73) | 9.81 (9.51 – 10.12) | 44.36 (43.72 – 45.01) |
|  | Not applicable | 866,690 | 9,507,665 | 44.99 (44.57 – 45.42) | 81.67 (81.10 – 82.25) | 299.83 (298.73 – 300.93) |
| Can-Marge: residential instability | Q1: lowest | 684,110 | 8,500,472 | 12.02 (11.79 – 12.26) | 20.77 (20.47 – 21.08) | 80.55 (79.95 – 81.16) |
|  | Q2 | 795,190 | 9,817,426 | 14.16 (13.92 – 14.39) | 24.29 (23.98 – 24.60) | 94.89 (94.29 – 95.50) |
|  | Q3 | 605,800 | 7,322,051 | 15.33 (15.05 – 15.62) | 27.21 (26.84 – 27.59) | 107.61 (106.86 – 108.36) |
|  | Q4 | 533,090 | 6,283,626 | 18.37 (18.03 – 18.70) | 32.31 (31.86 – 32.75) | 123.75 (122.89 – 124.63) |
|  | Q5: highest | 400,930 | 4,612,169 | 20.17 (19.77 – 20.59) | 35.68 (35.14 – 36.23) | 136.30 (135.24 – 137.37) |
| Can-Marge: ethnic concentration | Q1: lowest | 954,900 | 11,824,144 | 15.66 (15.43 – 15.88) | 27.14 (26.85 – 27.44) | 104.58 (104.00 – 105.16) |
|  | Q2 | 755,190 | 9,331,573 | 16.06 (15.80 – 16.32) | 27.83 (27.50 – 28.17) | 108.97 (108.31 – 109.64) |
|  | Q3 | 538,400 | 6,478,421 | 15.35 (15.05 – 15.66) | 27.04 (26.64 – 27.44) | 104.91 (104.13 – 105.70) |
|  | Q4 | 413,645 | 4,835,337 | 14.52 (14.18 – 14.86) | 25.65 (25.20 – 26.11) | 99.74 (98.85 – 100.63) |
|  | Q5: highest | 357,000 | 4,066,270 | 14.09 (13.73 – 14.46) | 25.08 (24.60 – 25.57) | 97.17 (96.22 – 98.14) |
| Can-Marge: material deprivation | Q1: lowest | 605,070 | 7,750,350 | 11.60 (11.36 – 11.84) | 21.02 (20.70 – 21.35) | 82.17 (81.53 – 82.81) |
|  | Q2 | 575,995 | 7,014,402 | 13.91 (13.64 – 14.19) | 24.66 (24.30 – 25.03) | 95.97 (95.25 – 96.70) |
|  | Q3 | 602,450 | 7,269,221 | 16.12 (15.83 – 16.41) | 28.30 (27.92 – 28.69) | 108.54 (107.78 – 109.30) |
|  | Q4 | 520,600 | 6,138,857 | 17.30 (16.98 – 17.63) | 30.04 (29.61 – 30.48) | 116.03 (115.18 – 116.89) |
|  | Q5: highest | 715,010 | 8,362,914 | 18.07 (17.78 – 18.36) | 30.58 (30.21 – 30.96) | 119.48 (118.74 – 120.22) |
| Can-Marge: dependence | Q1: lowest | 547,940 | 6,635,762 | 10.58 (10.34 – 10.83) | 18.20 (17.88 – 18.53) | 73.47 (72.82 – 74.12) |
|  | Q2 | 492,130 | 5,984,347 | 12.11 (11.83 – 12.39) | 21.07 (20.70 – 21.44) | 84.04 (83.31 – 84.78) |
|  | Q3 | 478,260 | 5,724,217 | 14.00 (13.70 – 14.31) | 24.25 (23.85 – 24.66) | 96.05 (95.25 – 96.86) |
|  | Q4 | 607,600 | 7,315,853 | 16.43 (16.13 – 16.72) | 28.86 (28.48 – 29.26) | 111.40 (110.64 – 112.17) |
|  | Q5: highest | 893,190 | 10,875,566 | 20.13 (19.86 – 20.40) | 35.40 (35.05 – 35.75) | 133.81 (133.12 – 134.49) |
| Urban form | Active urban core | 252,680 | 3,069,538 | 19.09 (18.61 – 19.59) | 33.77 (33.13 – 34.43) | 128.97 (127.71 – 130.25) |
|  | Transit-reliant suburb | 208,940 | 2,317,897 | 17.24 (16.72 – 17.79) | 30.13 (29.43 – 30.84) | 115.96 (114.58 – 117.35) |
|  | Car-reliant suburb | 1,281,505 | 15,625,226 | 13.35 (13.17 – 13.53) | 23.57 (23.57 – 24.05) | 95.30 (94.82 – 95.79) |
|  | Exurban | 172,455 | 2,161,801 | 11.77 (11.32 – 12.23) | 20.95 (20.35 – 21.57) | 84.26 (83.05 – 85.50) |
|  | Non-CMA/CA | 1,103,550 | 13,361,284 | 17.16 (16.94 – 17.38) | 29.27 (28.98 – 29.56) | 110.36 (109.80 – 110.92) |
| CMA/CA size | Population: >1,500,000 | 890,965 | 10,577,548 | 13.19 (12.98 – 13.41) | 23.83 (23.54 – 24.13) | 96.98 (96.39 – 97.58) |
|  | Population: 500,000-1,499,999 | 476,460 | 6,221,982 | 14.89 (14.59 – 15.19) | 25.61 (25.22 – 26.01) | 98.28 (97.50 – 99.06) |
|  | Population: 100,000-499,999 | 572,720 | 6,882,226 | 16.03 (15.73 – 16.33) | 28.53 (28.13 – 28.93) | 110.86 (110.08 – 111.65) |
|  | Population: 30,000-99,999 | 302,240 | 3,425,768 | 17.87 (17.43 – 18.32) | 30.02 (29.44 – 30.60) | 113.35 (112.23 – 114.48) |
|  | Population: 10,000-29,999 | 112,800 | 1,421,360 | 17.19 (16.52 – 17.88) | 30.18 (29.29 – 31.10) | 114.55 (112.80 – 116.32) |
|  | Non-CMA/CA | 663,950 | 8,006,861 | 16.71 (16.42 – 16.99) | 28.52 (28.15 – 28.89) | 107.29 (106.58 – 108.01) |
| Airshed | Western | 368,440 | 4,495,814 | 15.37 (15.01 – 15.74) | 29.56 (29.06 – 30.07) | 111.20 (110.23 – 112.18) |
|  | Prairie | 412,740 | 4,989,145 | 16.04 (15.69 – 16.39) | 26.93 (26.48 – 27.39) | 94.85 (94.00 – 95.71) |
|  | West Central | 175,410 | 2,080,580 | 18.78 (18.20 – 19.38) | 32.26 (31.50 – 33.04) | 118.27 (116.81 – 119.76) |
|  | Southern Atlantic | 280,320 | 3,418,800 | 16.16 (15.74 – 16.60) | 28.86 (28.29 – 29.43) | 111.86 (110.74 – 112.98) |
|  | East Central | 1,753,950 | 21,218,022 | 14.85 (14.69 – 15.02) | 25.58 (25.37 – 25.80) | 102.85 (102.42 – 103.28) |
|  | Northern | 28,265 | 333,382 | 9.99 (8.97 – 11.12) | 18.06 (16.67 – 19.56) | 79.94 (76.96 – 83.03) |

\* Number of participants rounded to the nearest 5. † Person-years rounded to the nearest whole number

We also included airshed as a covariate in our statistical models to control, at a larger geographic scale, for regional differences in mortality rates across Canada. Canada has six defined airsheds (i.e. Western, Prairie, West Central, East Central, Southern Atlantic, and Northern), and each is characterized by generally similar large-scale air masses and meteorological conditions.^28^

### Statistical methods

First, we applied the single-variable Cox proportional hazard model to assess the associations between mortality from IHD, CVD, or non-accidental causes and each selected covariate. This allowed us to demonstrate the need to control for these covariates in our primary analysis models. Then, we used the quantile g-computation model to calculate the hazard ratio (HR) and its 95% confidence interval (CI) for time to death attributable to IHD, CVD, or non-accidental causes per quartile increase in all four air pollutants (PM_2.5_, SO_2_, NO_2_, O_3_) from each specific sector. The quantile g-computation model was adjusted for age, sex, education level, income, occupational class, marital status, labor force status, visible minority, indigenous identity, immigrant status, Can-Marg indices, CMA/CA size, urban form, and airshed. In addition to the HR and its 95% CI for the mixture of the four air pollutants, the quantile g-computation model estimated the relative proportional contribution (i.e., weight) of each individual pollutant within the mixture on the overall health effect (i.e., occurrence of death).

The weight of an air pollutant exposure variable could be either positive or negative, indicating that exposure to this air pollutant positively or negatively contributed to death. All weights in one direction (i.e., positive or negative) are constrained to sum to 1. While the weights in a quantile g-computation model do not inherently indicate the magnitude of health effects or statistical significance of individual exposures, they do provide insight into the relative contribution of exposures within the same direction (positive or negative). For example, a large weight does not confirm that the exposure is significantly associated with the outcome on its own, but it may suggest that an exposure contributes more to the overall association with the outcome compared to others. Details regarding the quantile g-computation approach have been elaborated by Keil et al^10^.

We conducted sensitivity analyses stratifying by sex, since we observed higher risks of death from IHD and CVD in males than in females in our preliminary assessment. The quantile g-computation models were fitted by the “qgcomp” package ^29^ in the R statistical computing environment (version 4.2.3).

We calculated risk difference (*RD*) of death from IHD, CVD or non-accidental causes between the individuals exposed to a given quartile of the air pollutant mixture and those exposed to the reference level (i.e., the lowest quartile of the mixture) by Equation 1:

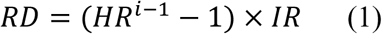

in which *HR* represents hazard ratio, *IR* denotes incidence rate of deaths from IHD, CVD, or non-accidental causes, and *i* is the *i*th quartile of the air pollutants.

## Results

We followed a cohort of 3,019,125 adults (48.3% male) across a total of 36,112,640 person-years (mean follow-up of 11.9 years). The mean age at cohort entry was 50.3 (standard deviation 15.4) years. Less than one quarter (23.6%) of the adults held university degrees, and 17.6% were immigrants. In our cohort, 42.4% of the population lived in car-reliant suburbs, and 0.9% were from the remote Northern airshed region. Over time, we identified 56,190 (60.7% male), 98,185 (54.8% male), and 381,050 (52.2% male) deaths from IHD, CVD, and non-accidental causes, corresponding to incidences of 1.56 (1.16), 2.72 (2.33), and 10.55 (9.56) deaths per 1000 person-years, respectively. Detailed socio-demographic characteristics of our cohort and incidence rates of deaths across different subgroups are shown in Table 1.

The interquartile ranges of exposure to ambient PM_2.5_, SO_2_, NO_2_, and O_3_ concentrations attributed to upstream petroleum sector emissions were 0.015 μg/m^3^, 0.004 ppb, 0.014 ppb, and 0.022 ppb, respectively, while those from residential fuel combustion were 0.124 μg/m^3^, 0.004 ppb, 0.522 ppb, and 0.000 ppb, respectively. Pollutant-and sector-specific exposure interquartile ranges and detailed distributions of population-weighted exposures are shown in Table S2 for all assessed sectors. The concentrations of the four air pollutants were inter-correlated across all five sectors. Especially, PM_2.5_ was strongly positively correlated with SO_2_ and NO_2_ in the upstream petroleum, downstream petroleum, and residential fuel combustion sectors. Conversely, O_3_ was negatively correlated with the other three air pollutants in the downstream petroleum and residential fuel combustion sectors (Figure S1).

The single-variable Cox models demonstrated that each of our selected covariates was significantly associated with mortality attributable to IHD, CVD, or non-accidental causes. For example, males appeared to have a higher risk of dying from IHD, CVD, or non-accidental causes compared to females. Additionally, the risk of dying decreased with higher levels of education and income. People living in non-CMA/CA areas or in the remote Northern airshed were more likely to die from IHD, CVD, or non-accidental causes than those living in CMA/CA areas or the East Central airshed. Statistics for other covariates are shown in Table S3.

The quantile g-computation models, which included all subjects, demonstrated significantly increased risks of non-accidental mortality associated with each quartile increase in exposure to the four air pollutants from residential fuel combustion (HR: 1.07, 95% CI: 1.05-, upstream petroleum (HR: 1.05, 95% CI: 1.04-1.05), downstream petroleum (HR: 1.03, 95% CI: 1.02-1.03), and the chemical industry (HR: 1.04, 95% CI: 1.04-1.05). These HRs and 95% CIs suggest that the risk differences of dying from non-accidental causes between individuals exposed to the highest quartile of these sector-specific air pollutants and individuals exposed to the lowest quartile are 12.07 (95% CI: 11.90-12.28) per 1000 person-years, 11.39 (95% CI: 11.20-11.63) per 1000 person-years, 12.00 (95% CI: 11.37-12.28) per 1000 person-years, and 12.96 (95% CI: 12.25-13.74) per 1000 person-years respectively. In these models, PM_2.5_ and/or SO_2_ consistently had the largest positive weights, suggesting that they contributed more significantly to the non-accidental mortality than NO_2_ and O_3_. Conversely, no clear association was observed for the non-ferrous smelting sector (HR: 1.01, 95% CI: 1.00-1.01).

Compared with non-accidental mortality, the positive associations of mortality from IHD with the mixtures of PM_2.5_, SO_2_, NO_2_, and O_3_ from the upstream petroleum, downstream petroleum, chemical industry, and residential fuel combustion sectors were stronger, as demonstrated by the larger HRs and their 95% CIs (see Figure 1). Specifically, the HRs (and their 95% CIs) for time to death attributable from IHD with each quartile increase in exposure to the four air pollutants from residential fuel combustion, upstream petroleum, downstream petroleum, the non-ferrous smelting industry, and the chemical industry were 1.18, (1.12–1.24), 1.12, (1.11–1.14), 1.06, (1.05–1.08), 1.02 (1.00–1.04), and 1.10, (1.08–1.13), respectively.

**Figure 1.**
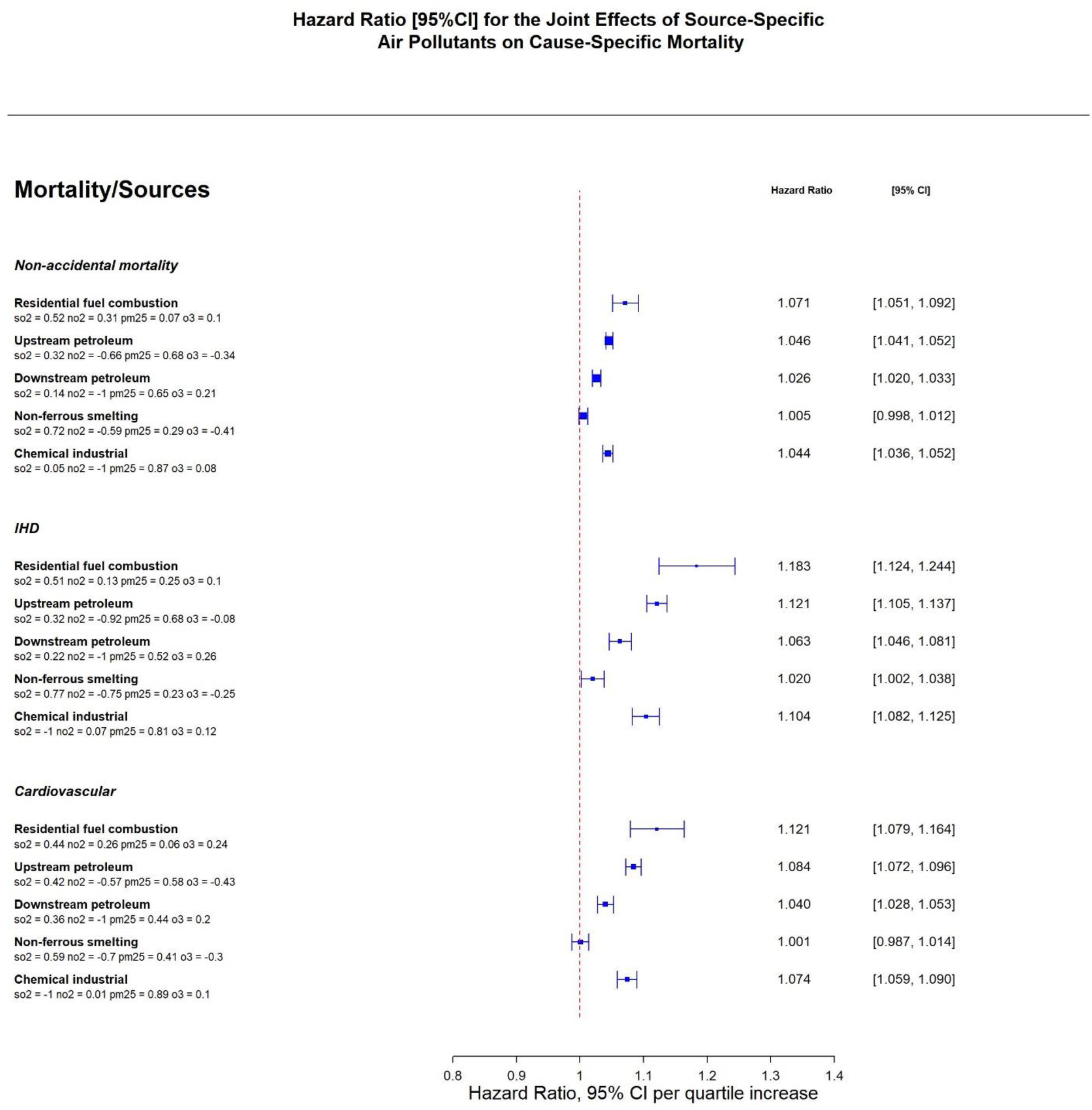
Adjusted HR (95% CI) and weights of the air pollutants from the quantile g-computation models for time to deaths attributable from ischemic heart disease (IHD), cardiovascular disease (CVD) and non-accidental causes per quartile increase in all the four air pollutants from one specific sector.

Similar increased risks of death were observed for CVD with each quartile increase in pollutant exposure from residential fuel combustion (HR: 1.12, 95% CI: 1.08–1.16), upstream petroleum (HR: 1.08, 95% CI: 1.07–1.10), downstream petroleum (HR: 1.04, 95% CI: 1.03–1.05), and the chemical industry (HR: 1.07, 95% CI: 1.06–1.09). In contrast, the non-ferrous smelting industry showed no significant associations with CVD (HR: 1.00, 95% CI: 0.99–1.01). Moreover, in the quantile g-computation models for the mortality from IHD or CVD, PM_2.5_ and SO_2_ always held larger weights in the positive direction than NO_2_ and O_3_, except in the mixture from the chemical industry, in which PM_2.5_ had a very large positive weight and SO_2_ showed a negative weight.

Similar results were observed in the analyses stratified by sex (Figure S2).

## Discussion

Our study revealed positive associations of mortality from IHD, CVD, and non-accidental causes with air pollution mixtures of PM_2.5_, SO_2_, NO_2_, and O_3_ from upstream and downstream petroleum, chemical industry, and residential fuel combustion. Moreover, our analyses, based on a large general population cohort, indicate that PM_2.5_ and SO_2_ are more important contributors than NO_2_ and O_3_ to the increased risk of the mortality from these causes.

These findings align with Canadian epidemiological studies investigating links between PM_2.5_ sulfur composition and adverse effects in children and adults.^30,31^ Korsiak et al^30^ found indications of stronger associations between PM_2.5_ and redox oxidant capacity (O_X_) air pollution and pediatric hospital admissions, notably when particulate sulfur and metal content was higher. The sulfate component of PM_2.5_ and ambient SO_2_ concentrations can be similarly influenced by sulfur emissions^32^, which could partly explain the higher contributions we observed with PM_2.5_ and SO_2_. Weichenthal et al^31^ also reported greater odds ratios for cardiovascular events in males when transition metal (e.g. copper) and sulfur particulate contents were above the median, compared to levels below the median. Internationally, a Northern China cohort study highlighted PM_2.5_ as a key contributor in their multi-pollutant models, with limited effects of NO_2_ due to attenuation from other pollutants.^33^ Our findings for petroleum sector-related health risk are consistent with source apportionment-based studies of source-specific CV or respiratory morbidity in the United States, which found significant positive associations for industry-related PM_2.5_ source types such as residual oil combustion^16,34^, associated with refining operations in the downstream petroleum sector, as well as with marine transportation in coastal and port areas.

Our findings for residential wood combustion are also consistent with prior findings of a positive association to CV morbidity for biomass combustion which included residential wood combustion.^15–18^ Our findings are consistent with mechanistic findings from exposure and toxicology studies. Controlled exposure and toxicological studies demonstrate that inhaled air pollutants are linked with oxidative stress, systemic inflammation, direct particle translocation, and autonomic nervous system imbalance.^35^ These pathways lead to physiological changes including endothelial dysfunction, thrombotic activity, epigenomic alterations, and the development of atherosclerosis and arrhythmogenesis.^35,36^ As these processes progress, conditions that may be subclinical like arrhythmias, hypertension, and arterial stiffness emerge.^35^ Over time, these subclinical changes manifest as clinical events, such as IHD, and ultimately contribute to CVD or other non-accidental mortality. These mechanisms are supported by numerous epidemiological studies associating long-term exposure to air pollutant(s) with increased risks of IHD, CVD, and/or other disease mortalities.^12,37–39^

Despite existence of them, most of the previous epidemiological studies have relied on single-pollutant or traditional multi-pollutant models, which is likely to encounter potential issues of collinearity. This can lead to misinterpretations, as the observed adverse health effect of a studied air pollutant may actually result from another concentration-highly-correlated air pollutant, rather than from its own toxicity. Additionally, while most previous studies examined only overall ambient (without disaggregating to contributing source types or sectors) or traffic-related air pollutants^12,40^, research on health effects of those from industrial or residential sectors is limited. Buteau et al^41^ separately assessed the association of industrial PM_2.5_ and SO_2_ with asthma onset in Quebec, Canada. Zhao et al^42^ explored associations between onset of systemic autoimmune rheumatic disease and the mixture of industrial PM_2.5_, SO_2_, and NO_2_ in Ontario, Canada. However, these two studies investigated air pollutants from all but not specific industrial sectors and only used distance to air pollution sources, tons of emitted air pollutants from the sources, and wind direction to roughly estimate air pollution exposures. Liu et al^43^ separately evaluated the associations of PM_2.5_, SO_2_, and NO_2_ from pulp and paper mills, petroleum refineries, and metal smelters with asthma onset, but did not pay attention to the actuality that concentrations of the three air pollutants from one of the industrial sectors were highly inter-correlated in space.

In contrast, we used the quantile g-computation model, that was developed specially for evaluating health effects of potentially inter-correlated exposures, to examine mixtures of air pollutants (PM_2.5_, SO_2_, NO_2_, and O_3_) from specific industrial or residential sectors. This approach not only avoids the occurrence of collinearity, but also enabled us to identify and prioritize sector-specific air pollutants contributing the most to IHD and CVD mortality risks. These findings can inform risk assessment and risk management by highlighting pollutants and sectors that may warrant targeted interventions to reduce mortality risks. Additionally, we employed the GEM-MACH model to more elaborately estimate exposures to the air pollutants from specific sectors by jointly considering atmospheric chemical processes and influences of meteorological factors and topography on the transport of air pollutants. Our findings revealed that mixtures from upstream and downstream petroleum, chemical industry, and residential fuel combustion were associated with IHD, CVD and non-accidental mortality, whereas mixtures from non-ferrous smelting was not associated with CVD and non-accidental mortality. This aligns with evidence that the chemical composition and associated health risk of ambient air pollutant exposures can vary by contributing source types or sectors.^8,9^ These findings suggest that targeting more toxic air pollutants (e.g., PM_2.5_ and SO_2_) from specific sectors may more efficiently reduce the disease burden attributable from long-term exposure to air pollution.

Building on these findings, our study also examined three sets of covariates to provide important context for understanding source-specific health risks within a broader social and geographic framework. The marginalization analyses align with prior research showing that individuals with lower socioeconomic status experience worse health outcomes, including lower life expectancy, higher mortality, and higher rates of chronic diseases like diabetes and asthma. ^44^ The urban form covariate demonstrated that denser, more active urban environments were associated with a lower risk of death from IHD, CVD, or non-accidental causes, whereas rural areas showed an increased risk. It is possible that the higher reliance on residential wood combustion in rural areas, which contributes to elevated levels of fine particulate matter (PM2.5) and other pollutants, may partially explain the observed increased risks in rural settings. These findings are consistent with previous studies that have found higher urban density is associated with lower cardiovascular disease risks. ^45,46^ Finally, the airshed analysis highlights important regional differences in health risks, with higher mortality HR for IHD, CVD and non-accidental causes in the Prairie, West Central, Southern Atlantic, and Northern airsheds. These regional variations in our findings underscore the importance of including smaller-scale geographically meaningful cohorts within a national cohort, particularly when these may reflect regional differences in source activity or meteorological patterns that may influence population exposures.

More strengths of this study include the use of the large, well-characterized CanCHEC cohort, which provides the statistical power necessary to detect small but significant changes in the health effects of ambient air pollution exposure. Additionally, our comprehensive adjustments for various covariates, including demographic, socioeconomic, marginalization index, environmental, and geographic factors, enhances the validity of our findings and their relevance to further analyses of health inequity and environmental justice.

Despite these strengths, there are several limitations to consider. While we accounted for many potential confounders, smoking information is not available in the CanCHEC cohort.

Cakmak et al^47^ used the Canadian Community Health Survey respondents from cycles between 2001 and 2011 to indirectly adjust observed HRs between lung cancer-related mortality and exposure to PM_2.5_ and O_3_ for smoking and found no significant influence of such indirect adjustment on the HRs. Weichenthal et al^48^ found minor effects of indirect adjustment for smoking on the HRs between exposure to ultrafine particles and incident brain tumors. Thus, we did not conduct any indirect adjustments for smoking in this study. Additionally, in one quantile g-computation model, we only considered exposure to multiple air pollutants from a single sector while in reality people are simultaneously exposed to multiple air pollutants from multiple sectors. Hence, an even more advanced statistical method is required to more closely simulate this real situation in the future.

In summary, our study demonstrates significant adverse associations between long-term exposure to multiple air pollutants from upstream and downstream petroleum, chemical industries, and residential fuel combustion, and increased risks of mortality from non-accidental causes, CVD, and IHD. We found that PM_2.5_ consistently had the largest contribution to elevated mortality risk, with additional contributions from SO₂. These findings suggest that air-pollution-attributable disease burden in Canada may be more effectively reduced by focusing air quality management efforts on the most toxic air pollutants from specific industrial and residential sources.

## Data Availability

Data is saved at Statistics Canada servers and can be accessed through a permission from Statistics Canada.

## Non-standard Abbreviations and Acronyms

CVD: Cardiovascular Diseases
IHD: Ischemic Heart Disease
PM_2.5_: Fine Particulate Matter
SO_2_: Sulfur Dioxide
NO_2_: Nitrogen Dioxide
O_3_: Ozone
HR: Hazard Ratio
CI: Confidence Interval
CanCHEC: Canadian Census Health and Environment Cohort
GEM-MACH: Global Environmental Multi-scale - Modelling Air Quality and Chemistry
NAPS: National Ambient Air Pollution Surveillance
DA: Dissemination Area
CAN-Marg: Canadian Marginalization Index
CMA/CA: Census Metropolitan Areas or Census Agglomerations
EPA: Environmental Protection Agency
WHO: World Health Organization
RD: Risk Difference
IR: Incidence Rate

## Acknowledgements

We would like to thank Statistics Canada for help accessing census and socioeconomic data, Environment and Climate change Canada (ECCC) for sector specific data, and Health Canada for the support for this publication.

---

**Figure S1.**
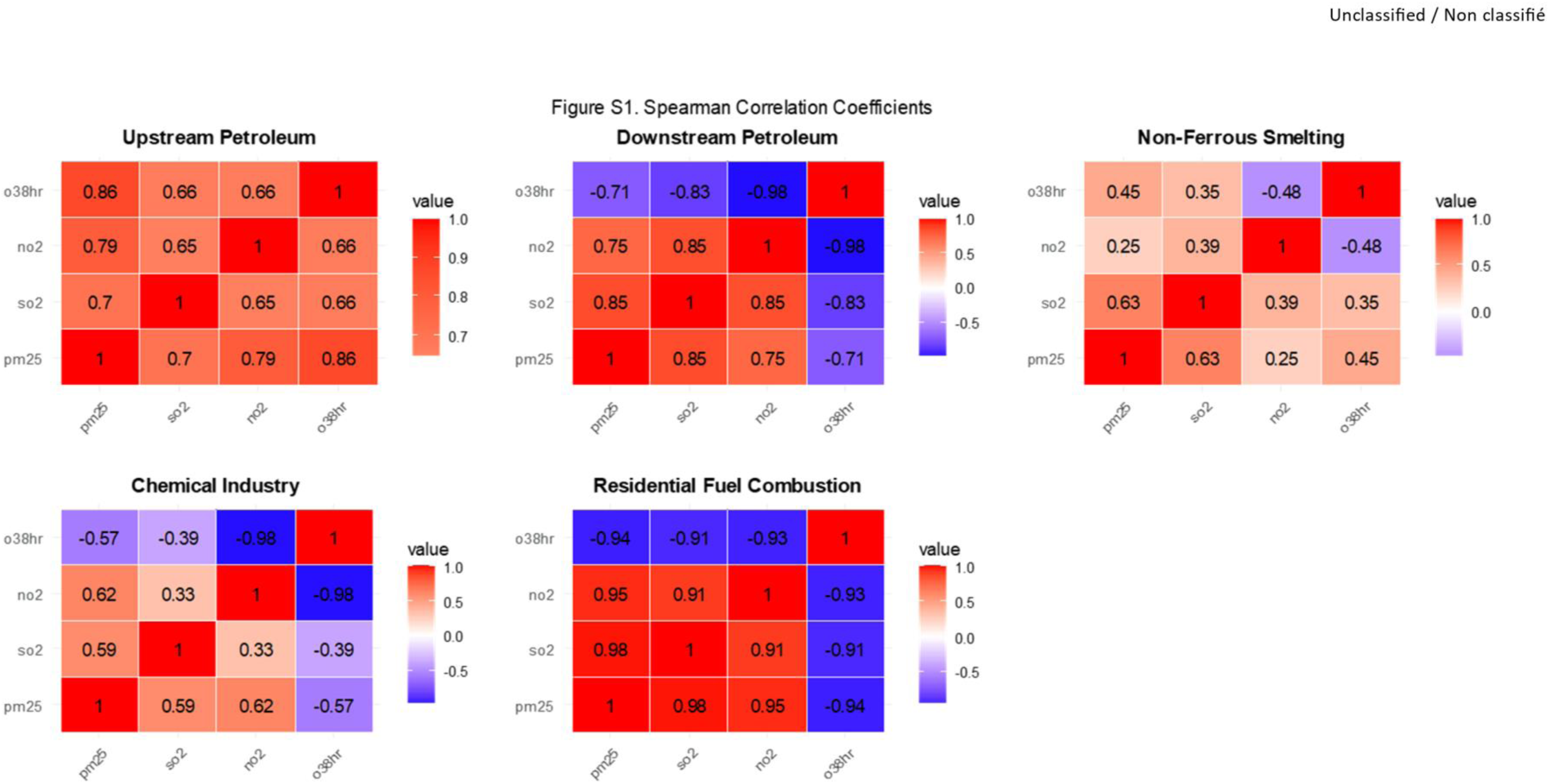
Spearman correlation coefficients of concentrations between any two of the air pollutants from upstream petroleum, downstream petroleum, non-ferrous smelting, chemical industry, and residential fuel combustion sectors.

**Figure S2.**
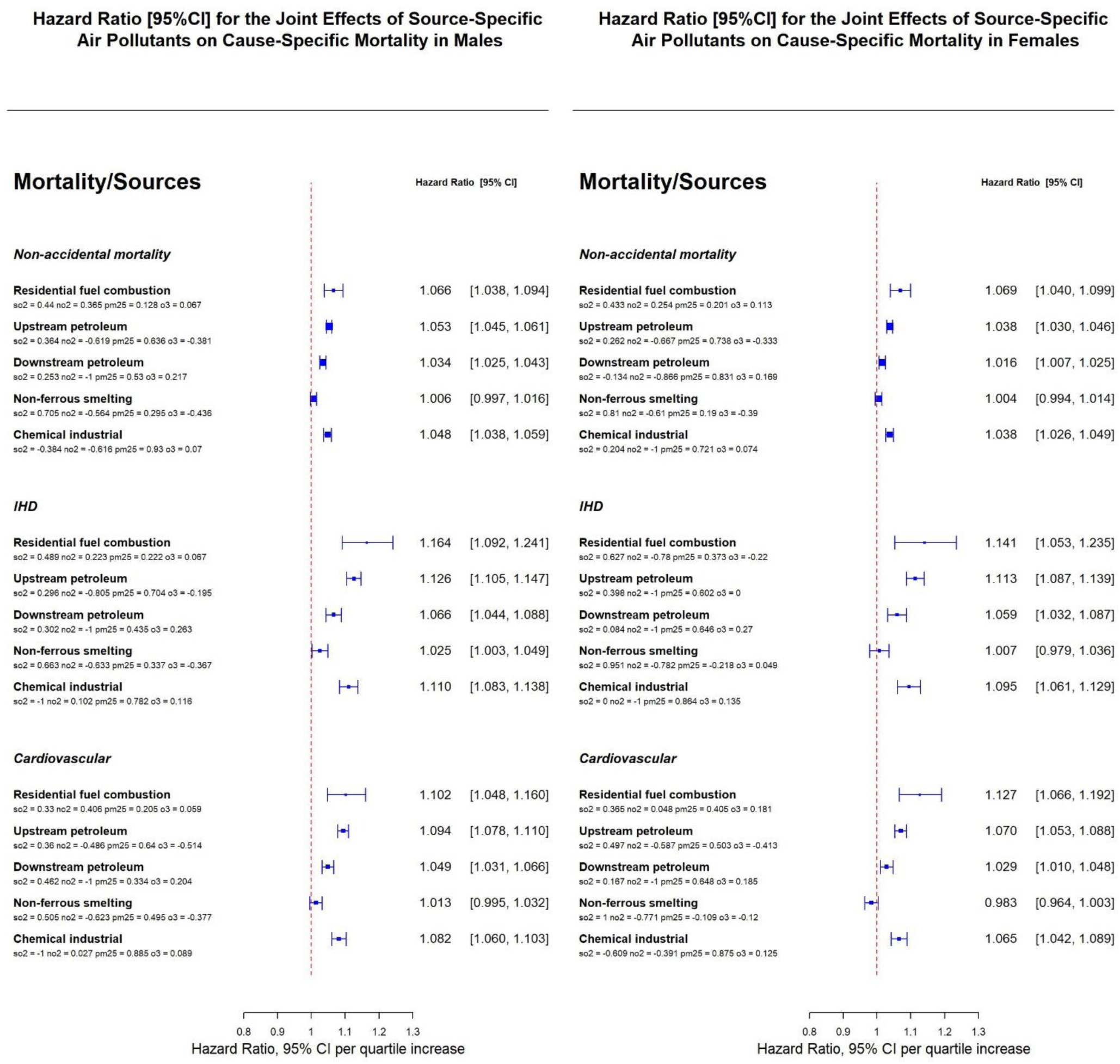
Adjusted HR (95% CI) and weights of the air pollutants from the quantile g-computation models for time to deaths attributable from ischemic heart disease (IHD), cardiovascular disease (CVD) and non-accidental causes per quartile increase in all the four air pollutants from one specific sector, using subpopulations of male (a) and female (b).

**Table S1.**
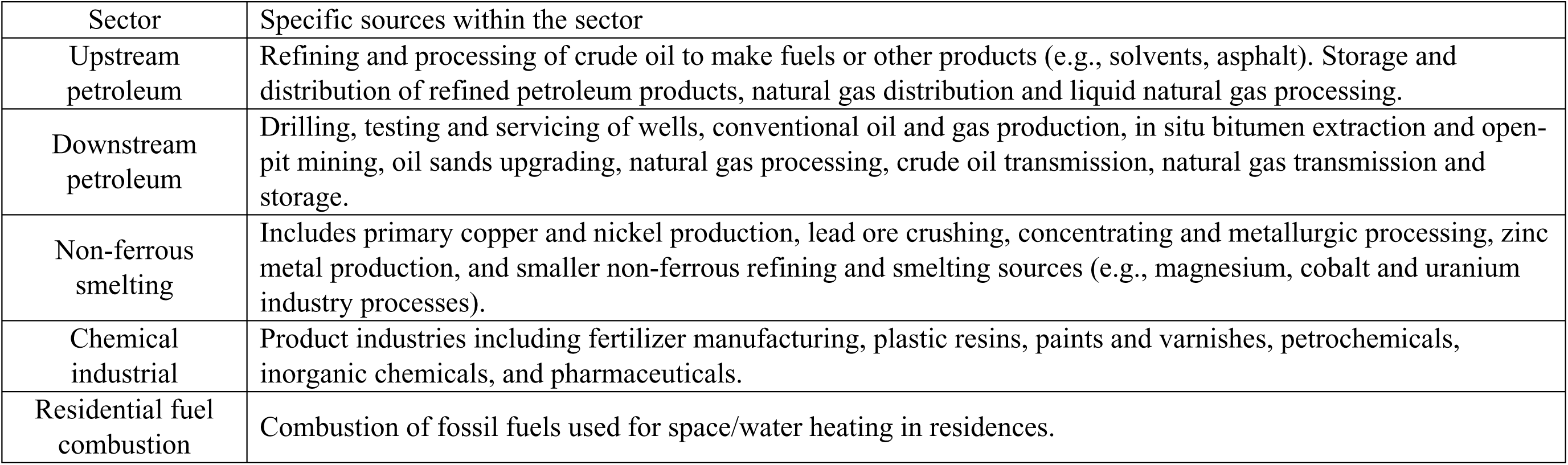
Descriptions of air pollutant emission inventory sectors.

**Table S2.**
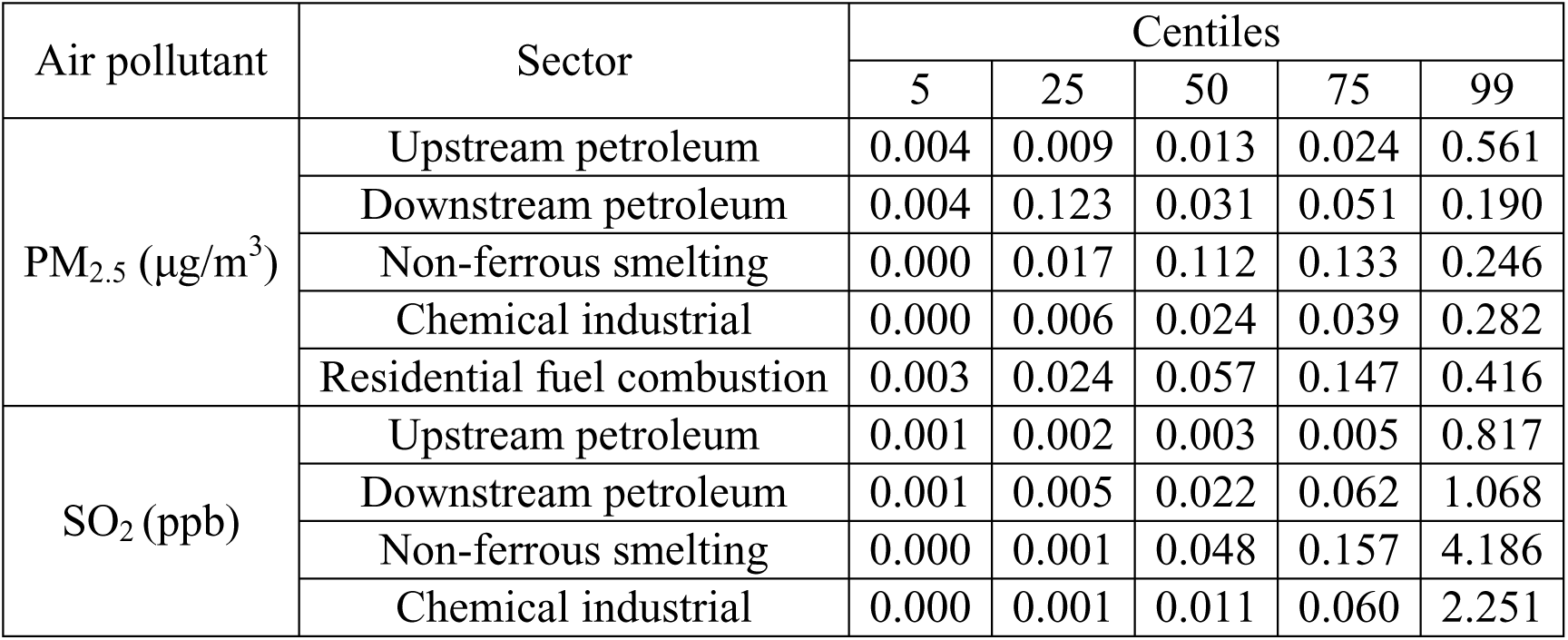
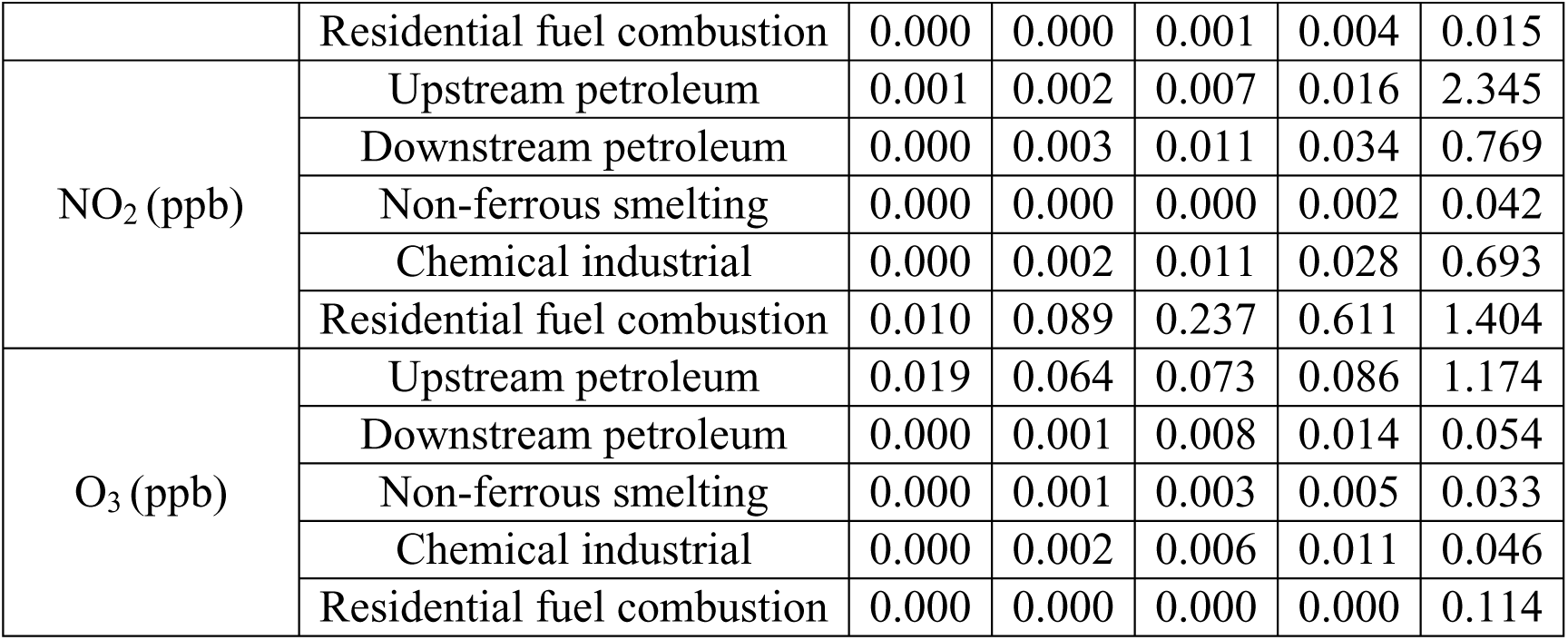
Distribution of population weighted concentrations of fine particulate matter (PM_2.5_), sulfur dioxide (SO_2_), nitrogen dioxide (NO_2_), and ozone (O_3_) from specific industrial or residential sectors.

| Air pollutant | Sector | Centiles |  |  |  |  |
| --- | --- | --- | --- | --- | --- | --- |
|  |  | 5 | 25 | 50 | 75 | 99 |
| PM<sub>2.5</sub> (µg/m<sup>3</sup>) | Upstream petroleum | 0.004 | 0.009 | 0.013 | 0.024 | 0.561 |
|  | Downstream petroleum | 0.004 | 0.123 | 0.031 | 0.051 | 0.190 |
|  | Non-ferrous smelting | 0.000 | 0.017 | 0.112 | 0.133 | 0.246 |
|  | Chemical industrial | 0.000 | 0.006 | 0.024 | 0.039 | 0.282 |
|  | Residential fuel combustion | 0.003 | 0.024 | 0.057 | 0.147 | 0.416 |
| SO<sub>2</sub> (ppb) | Upstream petroleum | 0.001 | 0.002 | 0.003 | 0.005 | 0.817 |
|  | Downstream petroleum | 0.001 | 0.005 | 0.022 | 0.062 | 1.068 |
|  | Non-ferrous smelting | 0.000 | 0.001 | 0.048 | 0.157 | 4.186 |
|  | Chemical industrial | 0.000 | 0.001 | 0.011 | 0.060 | 2.251 |
|  | Residential fuel combustion | 0.000 | 0.000 | 0.001 | 0.004 | 0.015 |
| NO<sub>2</sub> (ppb) | Upstream petroleum | 0.001 | 0.002 | 0.007 | 0.016 | 2.345 |
|  | Downstream petroleum | 0.000 | 0.003 | 0.011 | 0.034 | 0.769 |
|  | Non-ferrous smelting | 0.000 | 0.000 | 0.000 | 0.002 | 0.042 |
|  | Chemical industrial | 0.000 | 0.002 | 0.011 | 0.028 | 0.693 |
|  | Residential fuel combustion | 0.010 | 0.089 | 0.237 | 0.611 | 1.404 |
| O<sub>3</sub> (ppb) | Upstream petroleum | 0.019 | 0.064 | 0.073 | 0.086 | 1.174 |
|  | Downstream petroleum | 0.000 | 0.001 | 0.008 | 0.014 | 0.054 |
|  | Non-ferrous smelting | 0.000 | 0.001 | 0.003 | 0.005 | 0.033 |
|  | Chemical industrial | 0.000 | 0.002 | 0.006 | 0.011 | 0.046 |
|  | Residential fuel combustion | 0.000 | 0.000 | 0.000 | 0.000 | 0.114 |

**Table S3.** HR (95% CI) of each of the selected covariates for time to deaths attributable from ischemic heart disease (IHD), cardiovascular disease (CVD), or non-accidental causes from the single-variable Cox proportional hazard models.

| Covariate | subgroup | IHD | CVD | Non-accidental |
| --- | --- | --- | --- | --- |
| Sex | Female | Reference |  |  |
|  | Male | 2.09 (2.06-2.13) | 1.69 (1.67-1.71) | 1.45 (1.44-1.45) |
| Income quintile | Lowest | Reference |  |  |
|  | Lower-middle | 0.81 (0.79-0.82) | 0.83 (0.82-0.85) | 0.84 (0.83-0.84) |
|  | Middle | 0.70 (0.68-0.72) | 0.73 (0.72-0.74) | 0.74 (0.73-0.75) |
|  | Upper-middle | 0.60 (0.59-0.62) | 0.64 (0.63-0.66) | 0.65 (0.65-0.66) |
|  | Upper | 0.47 (0.46-0.49) | 0.52 (0.51-0.53) | 0.53 (0.53-0.54) |
| Education | <High school | Reference |  |  |
|  | High school | 0.84 (0.83-0.86) | 0.85 (0.84-0.86) | 0.85 (0.84-0.86) |
|  | Postsecondary non-university | 0.66 (0.64-0.68) | 0.70 (0.68-0.71) | 0.70 (0.69-0.71) |
|  | University degree | 0.56 (0.54-0.57) | 0.59 (0.58-0.60) | 0.59 (0.59-0.60) |
| Marital status | Common law |  |  |  |
|  | Married | 0.79 (0.76-0.82) | 0.86 (0.83-0.89) | 0.84 (0.82-0.85) |
|  | Never married/not common law | 1.30 (1.24-1.36) | 1.30 (1.25-1.35) | 1.24 (1.21-1.26) |
|  | Separated | 1.28 (1.20-1.36) | 1.31 (1.25-1.30) | 1.22 (1.18-1.23) |
|  | Divorced | 1.24 (1.18-1.30) | 1.25 (1.20-1.30) | 1.20 (1.18-1.23) |
|  | Widowed | 0.89 (0.85-0.93) | 0.99 (0.95-1.02) | 0.96 (0.94-0.97) |
| Employment status | Employed | Reference |  |  |
|  | Unemployed | 1.81 (1.69-1.93) | 1.83 (1.74-1.93) | 1.69 (1.64-1.73) |
|  | Not in labor force | 1.64 (1.60-1.69) | 1.69 (1.65-1.72) | 1.78 (1.76-1.79) |
| Immigrant | Yes | Reference |  |  |
|  | No | 1.30 (1.27-1.33) | 1.23 (1.21-1.25) | 1.28 (1.27-1.29) |
| Indigenous identity | No | Reference |  |  |
|  | Yes | 1.97 (1.89-2.05) | 1.93 (1.87-1.99) | 1.89 (1.86-1.92) |
| Visible minority | No | Reference |  |  |
|  | Yes | 0.95 (0.92-0.97) | 0.96 (0.94-0.98) | 0.95 (0.94-0.96) |
| Occupational | Management | Reference |  |  |
|  | Professional | 0.65 (0.58-0.72) | 0.66 (0.60-0.72) | 0.79 (0.76-0.82) |
|  | Skilled, technical, and supervisory | 1.08 (0.97-1.20) | 1.04 (0.96-1.13) | 1.10 (1.05-1.14) |
|  | Semi-skilled | 1.16 (1.04-1.29) | 1.12 (1.03-1.21) | 1.20 (1.15-1.25) |
|  | Unskilled | 1.33 (1.20-1.48) | 1.28 (1.18-1.40) | 1.33 (1.28-1.39) |
|  | Not applicable | 1.74 (1.57-1.92) | 1.71 (1.58-1.86) | 1.93 (1.85-2.00) |
| Can-Marge: residential instability | Q1: lowest | Reference |  |  |
|  | Q2 | 1.01 (0.99-1.04) | 1.00 (0.98-1.02) | 1.02 (1.01-1.03) |
|  | Q3 | 0.96 (0.93-0.98) | 0.97 (0.95-0.98) | 1.02 (1.01-1.03) |
|  | Q4 | 0.98 (0.95-1.01) | 0.97 (0.95-0.99) | 1.01 (1.00-1.02) |
|  | Q5: highest | 0.98 (0.95-1.01) | 0.96 (0.94-0.99) | 1.02 (1.01-1.04) |
| Can-Marge: ethnic concentration | Q1: lowest | Reference |  |  |
|  | Q2 | 0.98 (0.96-1.00) | 0.97 (0.96-0.99) | 1.00 (0.99-1.01) |
|  | Q3 | 0.96 (0.94-0.99) | 0.96 (0.95-0.98) | 0.99 (0.98-1.00) |
|  | Q4 | 0.90 (0.88-0.93) | 0.90 (0.89-0.92) | 0.94 (0.93-0.95) |
|  | Q5: highest | 0.85 (0.82-0.87) | 0.86 (0.84-0.88) | 0.88 (0.87-0.89) |
| Can-Marge: material deprivation | Q1: lowest | Reference |  |  |
|  | Q2 | 1.07 (1.04-1.10) | 1.04 (1.02-1.07) | 1.05 (1.04-1.06) |
|  | Q3 | 1.14 (1.11-1.18) | 1.10 (1.08-1.13) | 1.10 (1.09-1.11) |
|  | Q4 | 1.18 (1.15-1.21) | 1.12 (1.10-1.15) | 1.13 (1.12-1.14) |
|  | Q5: highest | 1.33 (1.29-1.36) | 1.24 (1.22-1.27) | 1.25 (1.24-1.27) |
| Can-Marge: dependence | Q1: lowest | Reference |  |  |
|  | Q2 | 0.93 (0.90-0.97) | 0.94 (0.92-0.96) | 0.94 (0.93-0.95) |
|  | Q3 | 0.94 (0.91-0.97) | 0.93 (0.91-0.95) | 0.94 (0.93-0.95) |
|  | Q4 | 0.94 (0.91-0.96) | 0.93 (0.91-0.96) | 0.94 (0.93-0.95) |
|  | Q5: highest | 0.94 (0.92-0.97) | 0.93 (0.91-0.95) | 0.93 (0.92-0.94) |
| Urban form | Active urban core | Reference |  |  |
|  | Transit-reliant suburb | 0.94 (0.90-0.97) | 0.93 (0.90-0.96) | 0.93 (0.91-0.94) |
|  | Car-reliant suburb | 0.87 (0.84-0.90) | 0.89 (0.87-0.91) | 0.90 (0.89-0.91) |
|  | Exurban | 0.95 (0.90-0.99) | 0.99 (0.96-1.03) | 0.97 (0.96-0.99) |
|  | Non-CMA/CA | 1.11 (1.08-1.14) | 1.10 (1.08-1.12) | 1.04 (1.02-1.05) |
| CMA/CA size | Non-CMA/CA | Reference |  |  |
|  | Population: 10,000-29,999 | 0.95 (0.91-0.99) | 0.96 (0.93-0.99) | 0.98 (0.96-1.00) |
|  | Population: 30,000-99,999 | 0.96 (0.93-0.99) | 0.94 (0.92-0.96) | 0.96 (0.95-0.97) |
|  | Population: 100,000-499,999 | 0.87 (0.85-0.90) | 0.90 (0.88-0.91) | 0.95 (0.94-0.96) |
|  | Population: 500,000-1,499,999 | 0.87 (0.85-0.90) | 0.87 (0.85-0.88) | 0.90 (0.89-0.91) |
|  | Population: >1,500,000 | 0.73 (0.71-0.75) | 0.76 (0.74-0.77) | 0.84 (0.83-0.85) |
| Airshed | East Central | Reference |  |  |
|  | Prairie | 1.25 (1.22-1.28) | 1.22 (1.20-1.24) | 1.06 (1.05-1.07) |
|  | West Central | 1.31 (1.27-1.36) | 1.31 (1.28-1.34) | 1.20 (1.18-1.22) |
|  | Southern Atlantic | 1.15 (1.12-1.18) | 1.20 (1.18-1.23) | 1.15 (1.13-1.16) |
|  | Western | 0.95 (0.93-0.98) | 1.06 (1.04-1.08) | 1.00 (0.99-1.01) |
|  | Northern | 1.15 (1.03-1.28) | 1.25 (1.16-1.36) | 1.29 (1.25-1.34) |

